# Sitting time enhances the effect of genetic liability to obesity on hypertension

**DOI:** 10.64898/2026.03.18.26348757

**Authors:** Chukwueloka Hezekiah, Daniel Bailey, Raha Pazoki

**Affiliations:** Cardiovascular and Metabolic Research Group, Department of Biosciences, College of Health, Medicine and Life Sciences, Brunel University of London, UB8 3PH, United Kingdom; Department of Public Health, Children’s, Learning Disability and Mental Health Nursing, Faculty of Health, Science, Social Care and Education, Kingston University, Kingston Hill, Kingston Upon Thames, Surrey, KT2 7LB, United Kingdom; Centre for Physical Activity in Health and Disease, College of Health, Medicine and Life Sciences, Brunel University of London, Uxbridge UB8 3PH, United Kingdom; Department of Sport, Health and Exercise Sciences, Brunel University of London, Uxbridge, UB8 3PH, United Kingdom; Department of Epidemiology and Biostatistics, School of Public Health, St Mary’s campus, Norfolk Place, Imperial College London, London W2 1PG, United Kingdom

**Author notes:** Corresponding author: Raha Pazoki, Cardiovascular and Metabolic Research Group, College of Health and Life Sciences, Brunel University of London, UB8 3PH, United Kingdom. Authors Email Address: Chukwueloka HEZEKIAH; Dr Daniel BAILEY.

**Keywords:** Hypertension, obesity, sedentary lifestyle, sitting time, genetic risk, UK Biobank

## Abstract

**Background and purpose:** Excessive sitting and genetic liability to obesity are associated with risk of obesity and hypertension, two significant risk factors for cardiovascular disease. This study aimed to investigate the interactive effects of genetic liability to obesity and excessive sitting on prevalence of hypertension.

**Methods:** Obesity genetic liability was estimated in unrelated individuals of European ancestry (n=208,594) using previously identified genetic variants and their effect sizes for adiposity related traits. Hypertension was defined as systolic blood pressure ≥ 140 mmHg, diastolic blood pressure ≥ 90 mmHg, or the use of anti-hypertensive medications. Logistic regression was used to examine the association between obesity genetic liability and across different levels of self-reported sitting time.

**Results:** excessive sitting and increased genetic liability were independently associated with higher odds of hypertension. The greatest odds of hypertension was observed in participants with high sitting time combined with increased genetic liability to obesity (OR=1.29; 95% CI = 1.25, 1.33, *P* <2 ×10^−16^) compared to individuals with low genetic liability and low sitting time. Interaction analysis identified that in individuals with excessive sitting, the effect of genetic liability of waist circumference on hypertension was greater compared to individuals with low sitting time (*P* _interaction_=0.03).

**Conclusion:** Combined excessive sitting and high genetic susceptibility to obesity is associated with greatest odds of hypertension. These findings highlight the importance of lifestyle in offsetting risk imposed by genetic factors.

## Introduction

The prevalence of hypertension is increasing globally, currently affecting an estimated 1.3 billion adults aged 30–79 years ^1^. This represents a 32% growth over the preceding two decades ^1^. Genetic variants (single nucleotide polymorphisms; SNPs) associated with obesity are strong predictors of hypertension risk ^2,3^. A mendelian randomisation study of two Swedish urban-based cohorts reported a causal link between genetically mediated adiposity and hypertension ^2^. In another study of over 300,000 UK Biobank (UKB) participants, higher genetic liability to obesity significantly increased the odds of prevalent hypertension ^4^. These findings underscore the importance of identifying factors to mitigate the risk of hypertension associated with genetic liability to obesity.

Sedentary behaviour, defined as low energy expenditure while sitting, reclining, or lying down during waking time ^5^, is a health risk behaviour associated with hypertension. A prospective observational study found that television (TV) viewing for over 3 hours per day was causally associated with an increased risk of hypertension ^6^. Furthermore, increasing daily sedentary behaviour is associated with higher odds of hypertension in a dose-response manner ^7,8^. Sedentary behaviour is also adversely associated with body mass index (BMI) and waist circumference (WC) ^9^. The relationship between sedentary activities (TV viewing, computer use, leisure screen time, and driving) and obesity has been shown to be causal ^10^, which may explain higher sedentary time in individuals with overweight and obesity compared with non-overweight individuals in the general population ^9,11^. As sedentary behaviour and obesity are each associated with hypertension, and causally associated with one another, it is plausible that they may interact to influence blood pressure. Examining how genetic predisposition to obesity may interact with sedentary behaviour to affect hypertension risk, therefore, requires investigation.

Behavioural and genetic factors can interact in complex ways, potentially attenuating or exacerbating health outcomes ^4^. Previous research has shown that individuals with a moderate or high genetic predisposition to obesity and low physical activity have increased odds of hypertension compared to individuals with low genetic liability to obesity and moderate or high physical activity levels ^4^. However, sedentary behaviour, which is distinct from physical inactivity ^5^, is independently associated with cardiometabolic health, including high blood pressure ^12^. While it is known that both genetic liability to obesity and higher sedentary behaviour are independently associated with increased risk of hypertension ^4^, the combined and interactive effects of these factors remain unexplored. Understanding this interaction could provide insights into personalised interventions for population groups at high risk of hypertension due to obesity-related genetic background and sedentary behaviour. The aim of this study, therefore, was to investigate the independent and interactive effects of genetic liability to obesity and sedentary time on the prevalence of hypertension in participants from the UKB.

## Methods

### Study population

The UKB is a large population-based prospective study to enable in-depth exploration of genetic and non-genetic factors contributing to disease. Over 500,000 individuals aged 40 to 69 years were enrolled into the UKB between 2006 and 2010 ^13^. After obtaining informed consent, the UKB collected various phenotype data during the initial assessment. This data, acquired through interviews and surveys, included socio-demographic, lifestyle, and health-related information. Additionally, participants underwent physical and anthropometric assessments and provided saliva, urine, and blood samples for proteomic, genetic, and metabolomic studies ^14^.

### Ethical approval

The Northwest Multi-Centre Research Ethics Committee (11/NW/0382) granted ethics approval to the UKB as a research tissue bank, and the UKB managed the ethical approval process. Ethical approval for analysing secondary data from the UKB in this present study was obtained from the College of Health, Medicine, and Life Sciences Research Ethics Committee at Brunel University of London (reference 27684-LR-Jan/2021-29901 1). Data for the research was sourced from the UKB using an approved data request application (ID 60549). The present study adhered to the Declaration of Helsinki principles ^15^.

### Genotyping and imputation

The UKB conducted the genotyping and imputation processes centrally, employing comprehensive techniques as detailed in previous publications ^16–18^. Participant blood samples were collected at the assessment centres, and their DNA was extracted and genotyped using the UKB Axiom Array designed to capture short insertions and deletions (indels) and SNPs ^16^. Genotype imputation was performed using three reference panels: the Haplotype Reference Consortium, UK10K, and 1000 Genomes phase 3, with the imputation carried out using the IMPUTE4 program. Additionally, the UKB calculated kinship coefficients and genetic principal components, which aided in identifying related individuals and accounting for population stratification ^16^.

### Sample for analysis

This study analysed data from a subset of unrelated European ancestry individuals from the UKB. The UKB collected genetic data from 488,377 individuals. After merging genetic data with phenotype data, 486,936 individuals were available for analysis. Excluded individuals were then those who withdrew consent (n=362), had first- or second-degree relatives in the dataset (n= 26,098), were of non-European ancestry (n= 27,117), had a sex mismatch (n= 328), were pregnant or uncertain about their pregnancy status at the start of the study (n= 513), did not disclose their smoking habits (n= 221), had missing pack-years of smoking data (n= 66,059), or were current or former smokers with zero pack-years of smoking calculated (n= 1049). Participants who were unsure about their time spent driving, using a computer during leisure time or TV viewing (n= 8681), or unsure about their dietary intake of fish, meat, fruits, or vegetables (n= 10,549) were also excluded. Additionally, participants who were taking cholesterol-lowering medication (n= 58,719), did not disclose their drinking habits (n= 132), had self-reported ancestry that did not match their genetic ancestry (n=42), or had missing data for the main study covariates (n= 78,834) were excluded. The final sample available for analysis after applying these criteria was 208,594 participants (**Supplementary Figure 1)**.

### Genetic liability to obesity

Genetic liability to obesity was estimated in the present study using previously identified genetic variants and their weights from genome-wide association studies (GWAS) for BMI (155 SNPs) ^19^, waist-hip ratio (WHR) (27 SNPs) and WC (41 SNPs) ^20^. As part of the SNP selection process, linkage disequilibrium (LD) pruning was assessed using the LDlink tool ^21^. Only SNPs that met the GWAS significance threshold of P < 5 × 10^−8^, SNPs with Minor allele frequency >0.01, and SNPs that did not exhibit dependence on another SNP based on the LD parameter of R^2^<0.1 were considered. The final list of SNPs used in the analyses is available in **Supplementary Table 1**. Plink v2.0 ^22,23^ was used to automatically calculate genetic liabilities by multiplying the effect estimate of each SNP related to obesity traits (BMI, WHR and WC) by the number of risk alleles each UKB participant carried at the corresponding SNPs. Plink then aggregates these products across all SNPs to generate an overall weighted genetic liability score for each obesity trait per individual. The outcome was standardised for analysis using the base package in R.

### Sedentary behaviour

During the initial UKB assessment, participants were asked to report the average time spent engaging in sedentary activities over a typical 24-hour day. This included any driving (e.g., car, bus, motorcycle, boat, truck), TV viewing and using a computer during leisure time. Participants who responded “Do not know” or “Prefer not to answer” to these questions were excluded. Following methodology from previous research ^24^, sedentary time for each participant was calculated as the sum of hours per day spent driving, TV viewing and using a computer during leisure time. Participants were categorised according to sedentary time. First, sedentary time was categorised as high and low using a median split to enhance statistical power. Tertiles of sedentary time (low, moderate and high) were also created to allow for more refined mapping of effect estimates across the spectrum of sedentary behaviour.

### Blood pressure and definition of hypertension

Blood pressure measurements were undertaken by the UKB centrally ^25^. Systolic blood pressure (SBP) and diastolic blood pressure (DBP) were each measured twice in mmHg using either an automated sphygmomanometer (Omron HEM 7015-T) or a manual sphygmomanometer if the automated reading could not be taken. The second reading was taken after a minimum interval of one minute following the first measurement. The automated measurements were taken with an appropriately sized cuff on each participant’s upper left arm (the right arm was used where it was not practical to use the left arm) while the participants were seated. For this study, the average of each the SBP and DBP measurement was used for analysis, which was calculated using the mean of these readings from the same type of device (manual or automated sphygmomanometer). Where two readings could not be taken using the same device, the mean of one automated and one manual measurement was used. For participants on blood pressure-lowering medication (n= 89,501), 15 mmHg was added to the SBP and 10 mmHg to the DBP reading, as suggested in previous studies ^4,26^. In line with American Heart Association guidelines ^27^, hypertension (stage 2) was defined as SBP ≥140 mmHg or DBP ≥90 mmHg, or the use of blood pressure-lowering medication.

### Study covariates

Covariates that may influence the outcome were identified in the UKB dataset. During the baseline UKB assessment, questionnaire data on dietary intake was collected including vegetables, fruits, fish and meat. Self-report physical activity was measured (walking, moderate and vigorous physical activity) to calculate weekly metabolic equivalent (MET) minutes. Smokers were categorised by UKB as current, past, or never smokers. The pack-years of smoking were calculated as: 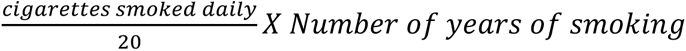 ^28^. Current or former smokers with any pack-year value were considered smokers, while participants who reported never smoking and had zero pack-years were classified as non-smokers ^4^. Additional covariates for the analysis were selected based on established cardiovascular disease risk factors commonly used in previous studies ^4,24^. These included alcohol consumption status (current, past, and non-drinkers) ^29^, low-density lipoprotein (LDL) cholesterol, high-density lipoprotein (HDL) cholesterol, and diabetes mellitus (defined as medically diagnosed diabetes, using insulin medication, serum haemoglobin A1c ≥ 48 mmol/mol [6.5%] or fasting glucose ≥ 7.0 mmol/dL ^30^).

### Statistical analysis

Using the *Mclust* and *Cluster* packages in R, a cluster analysis using the first 40 standardized genetic principal components was performed. The K-means algorithm ^31^ was used to calculate participants’ genetic distances, which requires specifying the number of clusters (K) ^32^. Based on the categories from the UKB self-reported phenotypic variable (ethnic background), a ‘K’ value of seven was used (**Supplementary Table 2**). A multidimensional scaling technique was applied to visualise similarities among individual clusters. Self-reported ancestry from the UKB phenotypic variable ‘ethnic background’ was compared with genetically inferred ancestry derived from cluster analysis, with particular attention given to the dominant clusters where the genetic ancestry was most prevalent. Participants whose self-reported ancestry did not match their genetically derived ancestry were excluded, as described above. Logistic regression was used to evaluate the odds of stage 2 hypertension with each unit increase in genetic liability to obesity as measured for BMI, WHR, and WC. Odds ratios (ORs) were calculated across three models: crude (Model 1), adjusted for age and sex (Model 2), and adjusted for age, sex, daily alcohol intake, smoking history, daily fruit and vegetable intake, meat and fish consumption, LDL, HDL, diabetes mellitus, and physical activity [MET-min/week] (Model 3).

The effect of genetic liability to obesity on hypertension was analysed for the whole sample and across sedentary time categories using logistic regression. The combined effect of genetic liability to obesity and sedentary time on hypertension was also assessed. For obesity genetic liabilities that demonstrated a statistically significantly association with hypertension in the whole sample, an interaction analysis was conducted to determine if sedentary time (high/low) modified the effect of genetic liability on hypertension. This interaction analysis was also undertaken for low, moderate and high (tertile-split) sedentary time categories. To quantify the independent effect of sedentary time on the observed associations, the modulatory effect of sedentary time on the association between genetic liability to obesity and hypertension was compared within sub-groups of similar genetic liabilities. Statistical significance was accepted when the *P*-value was <0.05. All statistical analyses were conducted using R version 4.0.0 ^33^.

## Results

The study included 208,594 participants (characteristics are shown in **Table 1**). The prevalence of hypertension was lower among participants with low sedentary time (42.5%) compared to high sedentary time (50.5%). Participants in the high sedentary time group had a significantly higher BMI.

**Table 1.** Characteristics of the study sample (n=208,594).

| Characteristic | Whole Sample<br>(n=208,594) | Sedentary time |  | P-value for difference in<br>sedentary time |
| --- | --- | --- | --- | --- |
|  |  | Low sedentary time<br>(≤4.5 hours/day)<br>(n=118,454) | High sedentary time<br>(>4.5 hours/day)<br>(n=90,140) |  |
| Age (Years) |  |  |  |  |
| Mean (SD) | 55.4 (8.0) | 55.1 (8.0) | 55.9 (8.0) | <0.001 |
| Median [Min, Max] | 56.0 [38, 73] | 56.0 [38, 73] | 57.0 [39, 72] |  |
| Sex |  |  |  |  |
| Female, n (%) | 115,179 (55.2%) | 73,269 (61.9%) | 41,910 (46.5%) | <0.001 |
| Male, n (%) | 93,415 (44.8%) | 45,185 (38.1%) | 48,230 (53.5%) |  |
| Stage 2 hypertension cases† |  |  |  |  |
| No, n (%) | 112,791 (54.1%) | 68,143 (57.5%) | 44,648 (49.5%) | <0.001 |
| Yes, n (%) | 95,803 (45.9%) | 50,311 (42.5%) | 45,492 (50.5%) |  |
| LDL (mmol/L) |  |  |  |  |
| Mean (SD) | 3.72 (0.81) | 3.68 (0.81) | 3.77 (0.81) | <0.001 |
| Median [Min, Max] | 3.67 [0.80, 9.74] | 3.63 [0.80, 9.74] | 3.73 [0.89, 9.04] |  |
| HDL (mmol/L) |  |  |  |  |
| Mean (SD) | 1.48 (0.38) | 1.53 (0.38) | 1.41 (0.36) | <0.001 |
| Median [Min, Max] | 1.43 [0.23, 4.40] | 1.49 [0.23, 4.40] | 1.36 [0.35, 4.13] |  |
| Smoking Status |  |  |  |  |
| Non-Smoker, n (%) | 138,211 (66.3%) | 83,042 (70.1%) | 55,169 (61.2%) | <0.001 |
| Smoker, n (%) | 70,383 (33.7%) | 35,412 (29.9%) | 34,971 (38.8%) |  |
| Systolic Blood Pressure<br>(mmHg) |  |  |  |  |
| Mean (SD) | 139 (20) | 137 (20) | 140 (20) | <0.001 |
| Median [Min, Max] | 137 [72, 268] | 135 [80, 253] | 139 [72, 268] |  |
| Diastolic Blood Pressure<br>(mmHg) |  |  |  |  |
| Mean (SD) | 83 (11) | 82 (11) | 85 (11) | <0.001 |
| Median [Min, Max] | 83 [42, 148] | 82 [45, 142] | 84 [42, 148] |  |
| Takes Blood Pressure<br>Lowering Medication |  |  |  |  |
| No, n (%) | 184,648 (88.5%) | 106,531 (89.9%) | 78,117 (86.7%) | <0.001 |
| Yes, n (%) | 23,946 (11.5%) | 11,923 (10.1%) | 12,023 (13.3%) |  |
| Alcohol Status |  |  |  |  |
| Never, n (%) | 6394 (3.1%) | 3827 (3.2%) | 2567 (2.8%) | <0.001 |
| Previous, n (%) | 6484 (3.1%) | 3464 (2.9%) | 3020 (3.4%) |  |
| Current, n (%) | 195,716 (93.8%) | 111,163 (93.8%) | 84,553 (93.8%) |  |
| Daily Fruit and Vegetable<br>Intake (n) |  |  |  |  |
| Mean (SD) | 7.88 (4.48) | 8.13 (4.48) | 7.56 (4.45) | <0.001 |
| Median [Min, Max] | 7.0 [0, 130] | 7.0 [0, 113] | 7.0 [0, 130] |  |
| Oily Fish Intake (n) |  |  |  |  |
| Mean (SD) | 1.63 (0.91) | 1.70 (0.90) | 1.60 (0.92) | <0.001 |
| Median [Min, Max] | 2.00 [0, 5] | 2.00 [0, 5] | 2.00 [0, 5] |  |
| Meat Intake (n) |  |  |  |  |
| Mean (SD) | 7.80 (2.79) | 7.60 (2.85) | 8.20 (2.68) | <0.001 |
| Median [Min, Max] | 8.00 [0, 25] | 8.00 [0, 20] | 8.00 [0, 25] |  |
| BMI (kg/m<sup>2</sup>) |  |  |  |  |
| Mean (SD) | 26.90 (4.54) | 26.10 (4.24) | 27.90 (4.72) | <0.001 |
| Median [Min, Max] | 26.30 [12.1, 66.2] | 25.50 [12.1, 63.6] | 27.30 [12.8, 66.2] |  |
| Diabetes Mellitus |  |  |  |  |
| No, n (%) | 202,445 (97.1%) | 115,512 (97.5%) | 86,933 (96.4%) | <0.001 |
| Yes, n (%) | 6149 (2.9%) | 2942 (2.5%) | 3207 (3.6%) |  |

Statistical analyses were performed using the chi-squared test for categorical variables and ANOVA for numerical variables. † Systolic blood pressure >140 mmHg or diastolic blood pressure >90 mmHg. LDL, low-density lipoprotein cholesterol; HDL, high-density lipoprotein cholesterol; BMI, body mass index.

The association of genetic liability to obesity and stage 2 hypertension across high/low sedentary time is shown in **Table 2**. For the whole sample, the odds of hypertension across all models were significantly higher for each unit increase in genetic liability for BMI, WHR and WC (OR=1.05, 1.03 and 1.04, respectively, for the most adjusted model) (**Table 2**). In participants with low sedentary time, the odds of hypertension were significantly higher for each unit increase in genetic liability to obesity across all models for BMI, WHR and WC (OR=1.04, 1.03 and 1.03, respectively, for the most adjusted model). Within the participant groups with high sedentary time, the odds of hypertension were also significantly greater with increasing genetic liability to obesity for BMI, WHR and WC models (OR= 1.05, 1.04 and 1.04, respectively) (**Table 2**). A significant interaction effect was found for genetic liability and sedentary time, where higher sedentary time increased the strength of association between WC genetic liability and hypertension across all models (*p* = 0.03; **Table 2**). This interaction was not significant for BMI or WHR genetic liabilities.

**Table 2.** Association between genetic liability to obesity and hypertension.

| Genetic Liability | Whole Sample<br>(n= 208,594) |  |  |  | Low sedentary time<br>(≤ 4.5 hours/day)<br>(n=118,454) |  |  | High sedentary time<br>(> 4.5 hours/day)<br>(n=90,140) |  |  |
| --- | --- | --- | --- | --- | --- | --- | --- | --- | --- | --- |
|  | Odds Ratio | 95% CI | *P*-value for Odds Ratio | Interaction *P*-value * | Odds Ratio | 95% CI | *P*-value for Odds Ratio | Odds Ratio | 95% CI | *P*-value for Odds Ratio |
| Model 1 |  |  |  |  |  |  |  |  |  |  |
| BMI | 1.04 | 1.03 - 1.05 | $2 \times 10^{-16}$ | 0.26 | 1.03 | 1.02 - 1.04 | $2.49 \times 10^{-6}$ | 1.05 | 1.03 - 1.06 | $2.16 \times 10^{-12}$ |
| WHR | 1.03 | 1.02 - 1.04 | $6.41 \times 10^{-12}$ | 0.53 | 1.03 | 1.02 - 1.04 | $5.68 \times 10^{-7}$ | 1.03 | 1.02 - 1.05 | $2.10 \times 10^{-6}$ |
| WC | 1.04 | 1.03 - 1.04 | $8.36 \times 10^{-14}$ | 0.06 | 1.03 | 1.02 - 1.04 | $9.50 \times 10^{-6}$ | 1.04 | 1.03 - 1.05 | $2.98 \times 10^{-8}$ |
| Model 2 <sup>a</sup> |  |  |  |  |  |  |  |  |  |  |
| BMI | 1.05 | 1.04 - 1.06 | $2 \times 10^{-16}$ | 0.24 | 1.04 | 1.02 - 1.05 | $1.08 \times 10^{-8}$ | 1.06 | 1.04 - 1.07 | $1.22 \times 10^{-14}$ |
| WHR | 1.04 | 1.03 - 1.04 | $1.40 \times 10^{-13}$ | 0.42 | 1.03 | 1.02 - 1.05 | $2.83 \times 10^{-7}$ | 1.04 | 1.02 - 1.05 | $9.47 \times 10^{-8}$ |
| WC | 1.04 | 1.03 - 1.04 | $1.48 \times 10^{-12}$ | 0.02 | 1.03 | 1.01 - 1.04 | $1.33 \times 10^{-4}$ | 1.04 | 1.03 - 1.06 | $4.59 \times 10^{-9}$ |
| Model 3 <sup>b</sup> |  |  |  |  |  |  |  |  |  |  |
| BMI | 1.05 | 1.04 - 1.06 | $2 \times 10^{-16}$ | 0.26 | 1.04 | 1.02 - 1.05 | $3.95 \times 10^{-8}$ | 1.05 | 1.04 - 1.07 | $9.10 \times 10^{-14}$ |
| WHR | 1.03 | 1.02 - 1.04 | $1.26 \times 10^{-11}$ | 0.44 | 1.03 | 1.02 - 1.04 | $2.67 \times 10^{-6}$ | 1.04 | 1.02 - 1.05 | $8.77 \times 10^{-7}$ |
| WC | 1.04 | 1.03 - 1.05 | $5.92 \times 10^{-13}$ | 0.03 | 1.03 | 1.01 - 1.04 | $4.05 \times 10^{-5}$ | 1.04 | 1.03 - 1.06 | $3.54 \times 10^{-9}$ |

Note: Odds ratios for stage 2 hypertension for each unit increase in standardised genetic liability.
Abbreviations: BMI, Body Mass Index; WHR, Waist Hip Ratio; WC, Waist Circumference; HbA1C, haemoglobin A1c.
<sup>a</sup> Adjusted for age and sex.
<sup>b</sup> Adjusted for age, sex, smoking status, alcohol status, meat and fish intake, fruit and vegetable intake, low-density lipoprotein cholesterol, high-density lipoprotein, diabetes (diagnosed by doctor or using insulin medication or glucose $\geq 7.0$ mmol/l or HbA1C $\geq 48$ mmol/mol (6.5%)), and physical activity (MET-min/week) as a continuous variable.
\* *P*-value is for the interaction between genetic liability for obesity and sedentary time (high or low) on an additive scale.

**Table 3** shows the combined effect of genetic liability and sedentary time on the odds of hypertension. Compared to the reference group (low sedentary time and low genetic liability), all other combinations of sedentary time and genetic liability were associated with increased odds of hypertension. Participants with high sedentary time and high genetic liability to obesity had the greatest odds of hypertension (most adjusted model: OR=1.29 for BMI, OR=1.25 for WHR, OR=1.29 for WC). This pattern was also reflected for prevalence of hypertension, which was markedly higher among participants with a combination of high sedentary time and high genetic liability to obesity (**Table 3**). Sedentary time was independently associated with hypertension as demonstrated by increased odds for high versus low sedentary time across all categories of genetic liability to obesity (**Table 4**). The greatest likelihood of hypertension was observed among participants with high sedentary time and high genetic liability.

**Table 3.** Prevalence and odds of stage 2 hypertension across sedentary time and genetic liability to obesity groups.

| Sedentary time group | Genetic Liability Categories |  | Hypertensive (n) | Non-Hypertensive (n) | Hypertension Prevalence (%) | Model 1 |  |  | Model 2<sup>a</sup> |  |  | Model 3<sup>b</sup> |  |  |
| --- | --- | --- | --- | --- | --- | --- | --- | --- | --- | --- | --- | --- | --- | --- |
|  |  |  |  |  |  | Odds Ratio | 95% CI | P-value for Odds Ratio | Odds Ratio | 95% CI | P-value for Odds Ratio | Odds Ratio | 95% CI | P-value for Odds Ratio |
| Low | BMI | Low | 16,824 | 23,584 | 41.64 | 1 (reference) |  |  | 1 (reference) |  |  | 1 (reference) |  |  |
| Low | | Moderate | 16,789 | 22,790 | 42.42 | 1.03 | 1.00 - 1.06 | $2.48 \times 10^{-2}$ | 1.04 | 1.01 - 1.07 | $1.40 \times 10^{-2}$ | 1.04 | 1.01 - 1.07 | $1.57 \times 10^{-2}$ |
| Low | | High | 16,698 | 21,769 | 43.41 | 1.08 | 1.05 - 1.11 | $4.77 \times 10^{-7}$ | 1.09 | 1.06 - 1.12 | $1.65 \times 10^{-8}$ | 1.09 | 1.06 - 1.12 | $5.12 \times 10^{-8}$ |
| High | | Low | 14,337 | 14,787 | 49.23 | 1.36 | 1.32 - 1.40 | $2 \times 10^{-16}$ | 1.22 | 1.18 - 1.26 | $2 \times 10^{-16}$ | 1.16 | 1.12 - 1.19 | $2 \times 10^{-16}$ |
| High | | Moderate | 15,110 | 14,842 | 50.45 | 1.43 | 1.39 - 1.47 | $2 \times 10^{-16}$ | 1.28 | 1.24 - 1.32 | $2 \times 10^{-16}$ | 1.22 | 1.18 - 1.26 | $2 \times 10^{-16}$ |
| High | | High | 16,045 | 15,019 | 51.65 | 1.50 | 1.45 - 1.54 | $2 \times 10^{-16}$ | 1.36 | 1.32 - 1.40 | $2 \times 10^{-16}$ | 1.29 | 1.25 - 1.33 | $2 \times 10^{-16}$ |
| Low | WHR | Low | 16,480 | 23,061 | 41.68 | 1 (reference) |  |  | 1 (reference) |  |  | 1 (reference) |  |  |
| Low | | Moderate | 16,882 | 22,709 | 42.64 | 1.04 | 1.01 - 1.07 | $6.10 \times 10^{-3}$ | 1.04 | 1.01 - 1.07 | $5.38 \times 10^{-3}$ | 1.04 | 1.01 - 1.07 | $1.41 \times 10^{-2}$ |
| Low | | High | 16,949 | 22,373 | 43.10 | 1.06 | 1.03 - 1.09 | $5.16 \times 10^{-5}$ | 1.07 | 1.03 - 1.10 | $3.02 \times 10^{-5}$ | 1.06 | 1.03 - 1.09 | $1.54 \times 10^{-4}$ |
| High | | Low | 14,840 | 15,156 | 49.47 | 1.37 | 1.33 - 1.41 | $2 \times 10^{-16}$ | 1.22 | 1.18 - 1.26 | $2 \times 10^{-16}$ | 1.16 | 1.12 - 1.20 | $2 \times 10^{-16}$ |
| High | | Moderate | 15,163 | 14,772 | 50.65 | 1.44 | 1.39 - 1.48 | $2 \times 10^{-16}$ | 1.30 | 1.25 - 1.34 | $2 \times 10^{-16}$ | 1.23 | 1.19 - 1.27 | $2 \times 10^{-16}$ |
| High | | High | 15,489 | 14,720 | 51.27 | 1.47 | 1.43 - 1.52 | $2 \times 10^{-16}$ | 1.32 | 1.28 - 1.37 | $2 \times 10^{-16}$ | 1.25 | 1.21 - 1.29 | $2 \times 10^{-16}$ |
| Low | WC | Low | 16,718 | 23,363 | 41.71 | 1 (reference) |  |  | 1 (reference) |  |  | 1 (reference) |  |  |
| Low | | Moderate | 16,780 | 22,534 | 42.68 | 1.04 | 1.01 - 1.07 | $5.59 \times 10^{-3}$ | 1.04 | 1.01 - 1.07 | $1.10 \times 10^{-2}$ | 1.04 | 1.01 - 1.07 | $6.87 \times 10^{-3}$ |
| Low | | High | 16,813 | 22,246 | 43.04 | 1.06 | 1.03 - 1.09 | $1.46 \times 10^{-4}$ | 1.05 | 1.02 - 1.09 | $4.67 \times 10^{-4}$ | 1.06 | 1.03 - 1.09 | $1.83 \times 10^{-4}$ |
| High | | Low | 14,566 | 14,900 | 49.43 | 1.37 | 1.33 - 1.41 | $2 \times 10^{-16}$ | 1.22 | 1.18 - 1.26 | $2 \times 10^{-16}$ | 1.16 | 1.12 - 1.20 | $2 \times 10^{-16}$ |
| High | | Moderate | 15,131 | 15,071 | 50.10 | 1.40 | 1.36 - 1.45 | $2 \times 10^{-16}$ | 1.25 | 1.21 - 1.29 | $2 \times 10^{-16}$ | 1.19 | 1.15 - 1.22 | $2 \times 10^{-16}$ |
| High | | High | 15,795 | 14,677 | 51.83 | 1.50 | 1.46 - 1.55 | $2 \times 10^{-16}$ | 1.36 | 1.31 - 1.40 | $2 \times 10^{-16}$ | 1.29 | 1.25 - 1.33 | $2 \times 10^{-16}$ |

Odds ratio, 95% confidence interval, and P-Value is for the joint effects of genetic liability for obesity and sedentary time (high or low) on hypertension. The values are derived from logistic regression models.
Abbreviations: BMI, Body Mass Index; WHR, Waist Hip Ratio; WC, Waist Circumference; HbA1C, haemoglobin A1c.
<sup>a</sup> Adjusted for age and sex.
<sup>b</sup> Adjusted for age, sex, smoking status, alcohol status, meat and fish intake, fruit and vegetable intake, low-density lipoprotein cholesterol, high-density lipoprotein, diabetes (diagnosed by doctor OR using insulin medication or glucose $\geq 7.0$ mmol/l or HbA1C $\geq 48$ mmol/mol (6.5%), and physical activity (MET-min/week) as a continuous variable.

**Table 4.**
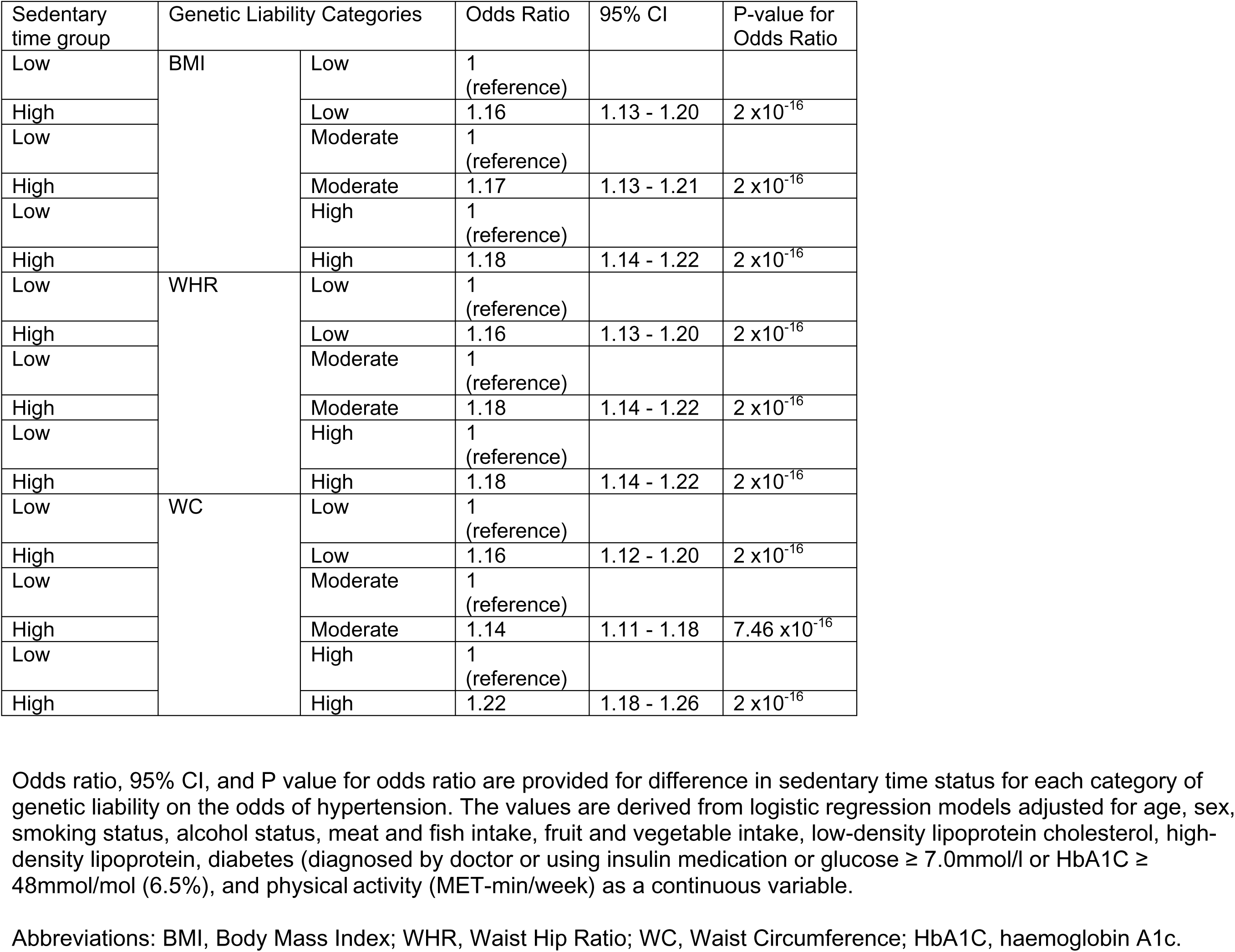
Odds of hypertension in high versus low sedentary time within categories of genetic liability to obesity.

| Sedentary time group | Genetic Liability Categories |  | Odds Ratio | 95% CI | P-value for Odds Ratio |
| --- | --- | --- | --- | --- | --- |
| Low | BMI | Low | 1 (reference) |  |  |
| High | | Low | 1.16 | 1.13 - 1.20 | $2 \times 10^{-16}$ |
| Low |  | Moderate | 1 (reference) |  |  |
| High | | Moderate | 1.17 | 1.13 - 1.21 | $2 \times 10^{-16}$ |
| Low |  | High | 1 (reference) |  |  |
| High | | High | 1.18 | 1.14 - 1.22 | $2 \times 10^{-16}$ |
| Low | WHR | Low | 1 (reference) |  |  |
| High | | Low | 1.16 | 1.13 - 1.20 | $2 \times 10^{-16}$ |
| Low |  | Moderate | 1 (reference) |  |  |
| High | | Moderate | 1.18 | 1.14 - 1.22 | $2 \times 10^{-16}$ |
| Low |  | High | 1 (reference) |  |  |
| High | | High | 1.18 | 1.14 - 1.22 | $2 \times 10^{-16}$ |
| Low | WC | Low | 1 (reference) |  |  |
| High | | Low | 1.16 | 1.12 - 1.20 | $2 \times 10^{-16}$ |
| Low |  | Moderate | 1 (reference) |  |  |
| High | | Moderate | 1.14 | 1.11 - 1.18 | $7.46 \times 10^{-16}$ |
| Low |  | High | 1 (reference) |  |  |
| High | | High | 1.22 | 1.18 - 1.26 | $2 \times 10^{-16}$ |

Odds ratio, 95% CI, and P value for odds ratio are provided for difference in sedentary time status for each category of genetic liability on the odds of hypertension. The values are derived from logistic regression models adjusted for age, sex, smoking status, alcohol status, meat and fish intake, fruit and vegetable intake, low-density lipoprotein cholesterol, high-density lipoprotein, diabetes (diagnosed by doctor or using insulin medication or glucose $\geq 7.0$ mmol/l or HbA1C $\geq 48$ mmol/mol (6.5%)), and physical activity (MET-min/week) as a continuous variable.
Abbreviations: BMI, Body Mass Index; WHR, Waist Hip Ratio; WC, Waist Circumference; HbA1C, haemoglobin A1c.

When sedentary time was stratified by tertiles, the odds of hypertension across all models were significantly higher for each unit increase in genetic liability for obesity within the whole sample (most adjusted model: OR = 1.05 for BMI, 1.03 for WHR, and 1.04 for WC; **Supplementary Table 3**).This pattern was also observed in participants with low (most adjusted model: OR = 1.03 for BMI, 1.03 for WHR, and 1.03 for WC), moderate (most adjusted model: OR = 1.04 for BMI, 1.04 for WHR, and 1.04 for WC) and high (most adjusted model: OR = 1.05 for BMI, 1.03 for WHR, and 1.04 for WC) sedentary time; **Supplementary Table 3**). No significant interaction effects for genetic liability and sedentary time were observed. The results for the combined effect of genetic liability and sedentary time on the odds of hypertension were largely consistent with sedentary time stratified by a median split. Other than moderate WHR genetic liability and low sedentary time, all combinations of genetic liabilities (BMI, WHR, and WC) and sedentary time increased the odds of hypertension compared with the reference group (low genetic liability and low sedentary time) (**Supplementary Table 4**).

## Discussion

The main findings of this study demonstrate, for the first time, that higher sedentary time is associated with hypertension across all levels of genetic liability to obesity. A combination of high genetic liability to obesity and high sedentary time presented the greatest risk, therefore highlighting a group that needs particular focus for prevention and management of hypertension. The current study supports previous findings that genetic liability to obesity increases the risk of hypertension ^4^. A mendelian randomisation study demonstrated this association to be causal ^2^, reinforcing the notion that genetic predisposition to obesity directly contributes to elevated blood pressure. This underscores the importance of identifying individuals with high genetic liability to obesity for targeted hypertension prevention strategies.

This is the first study to explore the combined effects of genetic liability to obesity and sedentary time on hypertension. Individuals with high genetic predisposition to obesity and high sedentary time had the greatest likelihood of hypertension. The exact mechanisms through which this combination of genetic and behavioural factors infers higher risk of hypertension remains to be elucidated. It is possible that high sedentary time disturbs energy homeostasis, which may promote fat accumulation, particularly visceral fat, that is metabolically active and pro-hypertensive ^10^. Informed by the present findings, reductions in sedentary time may be especially important in individuals genetically predisposed to obesity. This study contributes to the advancement of precision medicine by identifying individuals with genetic variants associated with obesity and high sedentary time as a population group that may benefit from targeted strategies for prevention and management of hypertension. Healthcare professionals may incorporate genetic liability assessments into routine care to identify individuals who may benefit most from intervention. Such targeted approaches could offer a novel approach for reducing hypertension prevalence and related complications (e.g., stroke) in individuals most at need, leading to more effective interventions and reduced burden on healthcare systems.

We previously reported that moderate and high levels of physical activity attenuated the odds of hypertension associated with high genetic liability to BMI in over 230,000 European ancestry participants from the UKB ^4^. The associations of sedentary time with hypertension across different levels of genetic liability to obesity in the present study remained after adjusting for physical activity, demonstrating that sedentary time is associated with hypertension regardless of physical activity level. Moreover, the effect of sedentary time was consistent across strata of genetic liability to obesity, indicating that its adverse impact is independent of genetic predisposition. These findings suggest that sedentary time be considered an independent risk factor for hypertension regardless of individuals’ genetic predisposition to obesity and physical activity level.

Another key novel finding was the significant interaction effect demonstrating that high sedentary time exacerbated the effects of WC genetic liability on hypertension. Interaction effects were not observed for genetic liabilities related to BMI or WHR, which may reflect their phenotypic expressions having less association with visceral fat compared to WC ^34^. Waist circumference is a direct proxy for visceral adipose tissue ^35^, which is associated with an increased risk of hypertension ^36^. It is plausible that low energy expenditure inherent of higher volumes of sedentary time may exacerbate the association between WC genetic liability and hypertension through visceral fat accumulation. This study highlights the importance of identifying individuals with genetic variants associated with WC and supporting them with interventions to limit sedentary time.

The present study emphasises the significance of limiting sedentary time for reducing hypertension risk across different levels of genetic predisposition to obesity. Interventions targeting reductions in sitting have been effective for lowering blood pressure ^37^. For example, the workplace ‘I-STAND Intervention’, involving health coaching, sitting reduction goals, a fitness tracker and a standing desk, reduced sitting by 31 minutes/day after six months, leading to a 3.48 mmHg reduction in systolic blood pressure ^38^. The synergistic effect of genetic liability to obesity and sedentary time suggests that interventions may be particularly important for individuals genetically predisposed to obesity, but interventions targeting specific genetic groups are yet to be explored and may explain why some interventions are ineffective ^39^. Researchers and clinicians could use genetic screening during routine blood tests to identify individuals with high genetic liability to obesity and prioritise them for personalised blood pressure management interventions focused on sedentary behaviour. Targeted interventions that integrate behavioural strategies, such as sedentary behaviour, with genetic liability profiling may offer a more effective approach to reducing hypertension than interventions delivered to individuals irrespective of their genetic predisposition; this should be investigated in future studies.

The main strengths of this study include the large sample size, which provided sufficient statistical power to explore the interactive relationship between obesity genetic factors and sedentary time on the prevalence of hypertension. Additionally, extensive phenotyping within the UKB enabled rigorous adjustment for confounding variables, enhancing precision of the estimated odds ratios in this study. The various subgroup analyses enabled quantification of the modulatory effect of sedentary time on the association between genetic liability to obesity and hypertension, enabling identification of phenotypes that present with the highest likelihood of hypertension.

A limitation of this study is the use of self-report data to measure sedentary time, which might bias outcomes due to misreporting ^40^. Future studies using device-based measures may provide more precise estimates of the effects observed. Another limitation is that the UKB cohort is not representative of the general population, tending to be healthier overall ^41^. This may introduce a ‘healthy volunteer’ bias in the observed associations ^42^.

## Conclusion

This study demonstrates that higher sedentary time is associated with hypertension across all levels of genetic predisposition to obesity, with this association being strongest in individuals with high genetic liability. Higher sedentary time exacerbates the effects of WC genetic liability on hypertension. These findings have identified population phenotypes that may benefit from targeted interventions. Further research is needed to evaluate the effectiveness of targeted sedentary behaviour interventions aimed at reducing hypertension risk in individuals genetically susceptible to obesity.

## Author Contributions

Conceptualization, R.P.; Data curation, C.H.; Formal analysis, C.H.; Funding acquisition, R.P.; Investigation, C.H.; Methodology, C.H., D.P.B. and R.P.; Project administration, R.P.; Resources, R.P.; Supervision, R.P. and D.P.B; Writing—original draft, C.H.; Writing—review & editing, C.H. D.P.B. and R.P. All authors have read and agreed to the published version of the manuscript.

## Funding and UKB application

UKB genotyping was supported by the British Heart Foundation (grant SP/13/2/30111) for Large-scale comprehensive genotyping of UKB for cardiometabolic traits and diseases: UK CardioMetabolic Consortium.

## Institutional Review Board Statement

The study was conducted in accordance with the Declaration of Helsinki and approved by the Institutional Review Board (or Ethics Committee) of Brunel University London, College of Health, Medicine and Life Sciences (27684-LR-Jan/2021-29901-1).

## Informed Consent Statement

Informed consent was obtained from all subjects involved in the study.

## Data Availability Statement

Not applicable.

## Acknowledgments

This study has been performed using the UKB application 60549.

